# Population-level COVID-19 mortality risk for non-elderly individuals overall and for non-elderly individuals without underlying diseases in pandemic epicenters

**DOI:** 10.1101/2020.04.05.20054361

**Authors:** John P.A. Ioannidis, Cathrine Axfors, Despina G. Contopoulos-Ioannidis

## Abstract

**OBJECTIVE:** To provide estimates of the relative risk of COVID-19 death in people <65 years old versus older individuals in the general population, the absolute risk of COVID-19 death at the population level during the first epidemic wave, and the proportion of COVID-19 deaths in non-elderly people without underlying diseases in epicenters of the pandemic.

**ELIGIBLE DATA:** Countries and US states with at least 800 COVID-19 deaths as of April 24, 2020 and with information on the number of deaths in people with age <65. Data were available for 11 European countries (Belgium, France, Germany, Ireland, Italy, Netherlands, Portugal, Spain, Sweden, Switzerland, UK), Canada, and 12 US states (California, Connecticut, Florida, Georgia, Illinois, Indiana, Louisiana, Maryland, Massachusetts, Michigan, New Jersey and New York) We also examined available data on COVID-19 deaths in people with age <65 and no underlying diseases.

**MAIN OUTCOME MEASURES:** Proportion of COVID-19 deaths in people <65 years old; relative risk of COVID-19 death in people <65 versus ≥65 years old; absolute risk of COVID-19 death in people <65 and in those ≥80 years old in the general population as of May 1, 2020; absolute COVID-19 death risk expressed as equivalent of death risk from driving a motor vehicle.

**RESULTS:** Individuals with age <65 account for 4.8-9.3% of all COVID-19 deaths in 10 European countries and Canada, 13.0% in the UK, and 7.8-23.9% in the US locations. People <65 years old had 36- to 84-fold lower risk of COVID-19 death than those ≥65 years old in 10 European countries and Canada and 14- to 56-fold lower risk in UK and US locations. The absolute risk of COVID-19 death as of May 1, 2020 for people <65 years old ranged from 6 (Canada) to 249 per million (New York City). The absolute risk of COVID-19 death for people ≥80 years old ranged from 0.3 (Florida) to 10.6 per thousand (New York). The COVID-19 death risk in people <65 years old during the period of fatalities from the epidemic was equivalent to the death risk from driving between 13 and 101 miles per day for 11 countries and 6 states, and was higher (equivalent to the death risk from driving 143-668 miles per day) for 6 other states and the UK. People <65 years old without underlying predisposing conditions accounted for only 0.7-2.6% of all COVID-19 deaths (data available from France, Italy, Netherlands, Sweden, Georgia, and New York City).

**CONCLUSIONS:** People <65 years old have very small risks of COVID-19 death even in pandemic epicenters and deaths for people <65 years without underlying predisposing conditions are remarkably uncommon. Strategies focusing specifically on protecting high-risk elderly individuals should be considered in managing the pandemic.

## INTRODUCTION

As the coronavirus disease 2019 (COVID-19) pandemic has spread widely around the globe,^1,2^ estimates about its eventual impact in terms of total number of deaths have varied widely, as they are mostly based on mathematical models with various speculative assumptions. It is crucial to estimate how much smaller the risk of death is among non-elderly people (<65 years old) as opposed to older individuals and how frequent deaths are in people who are <65 years old and have no underlying predisposing diseases. Media have capitalized on stories of young healthy individuals with severe, fatal outcomes. However, exaggeration should be avoided in responding to the pandemic.^3^ Accurate estimates of death risk at different age groups have important implications. Deaths of young, healthy people contribute far more quality-adjusted life-years lost than deaths in elderly individuals with pre-existing morbidity. Knowledge of COVID-19 death risks for people <65 years old at the population level can help guide different management strategies for the pandemic. People <65 years old represent the lion’s share of the workforce.

Here, we used data from 11 European countries, Canada and 12 states in the USA that are epicenters of the pandemic with a large number of deaths and where data were available for deaths according to age stratification. We aimed to evaluate the relative risk of death in people <65 years old versus older individuals in the general population, to provide estimates of absolute risk of COVID-19 death in these epicenters during the first epidemic wave, and to understand what proportion of COVID-19 deaths occur in people <65 years old and without underlying diseases.

## METHODS

We considered data from publicly reported situational reports of countries and US states or major cities that have been major epicenters of the pandemic as of late April. Eligibility criteria included: (1) at least 800 deaths accumulated as of April 24, 2020 (so as to qualify for a hotbed of the epidemic and to have a meaningful amount of data to analyze); and (2) information available on death counts per age strata, allowing to calculate numbers of deaths in people with age <65 or, alternatively, at least in people with age <60.

For each of the eligible geographical locations, we extracted information from the most up-to-date situational reports on April 24, 2020 focusing on total number of deaths with available age stratification, number of deaths in age <65 (or, if not available, number of deaths in age <60 and in age 60-69), number of deaths in age ≥80 (or, if not available, number of deaths in age ≥75) and, correspondingly, the proportions of the total deaths in each of these age categories. Information was extracted independently in duplicate by two authors (JI, CA) and discrepancies were resolved. Whenever information was unavailable for the desirable <65 years cut-off, we contacted the respective authorities issuing the situational report. We also extracted information on the proportion of men for all deaths and for deaths in each of these age categories, whenever available. For secondary analyses, we also extracted information on deaths in the subgroups of age <40 and age 40-64, whenever available (or, if not available, on <45 and 45-64).

One author (DC-I) downloaded information on the proportion of the population in each eligible location for each age group. We used census information from populationpyramid.net/world/2019 for countries and from worldpopulationreview.com for the US states.

We calculated the population-level relative risk of COVID-19 death for an individual <65 years old as compared with an individual ≥65 years old for each eligible country and US state/city. This is calculated as the ratio of (COVID-19 deaths with age <65 /population with age <65 in the respective age-pyramid) divided by (COVID-19 deaths with age ≥65/population with age ≥65 in the respective age-pyramid). Inverting this relative risk shows how many fold lower the risk of COVID-19 death is for an individual <65 years old as compared with an individual ≥65 years old.

We also calculated the absolute risk of dying with COVID-19 during the first epidemic wave for a person <65 years old in each eligible country and state by dividing the number of COVID-19 deaths (updated as of the end of day May 1, 2020) in this age group by the census population in this age group. Certainly, the number of deaths will increase and there is some uncertainty about the total projected number of deaths in each of these locations when this epidemic wave has passed, and all deaths have been counted. Most, if not all, locations seem to have passed the peak of the death curve as of May 1 and many are even close to resolution of the wave. However, we plan to update these numbers of absolute risk when the complete wave has passed. In order to get an estimate of the peak death risk during the epidemic period where fatalities occur, we also documented for each country and state the peak number of deaths from a 7-day moving frame up to May 1, 2020. This provides the number of deaths per day during the week of peak mortality risk. We used a 7-day moving frame, because there is some unavoidable noise fluctuation in death counts every day, plus for several countries and states there may be worse reporting delays for deaths during the weekend days. We used the actual date that deaths occurred for these estimates, but, whenever this was unavailable, we used the date of death reporting.

The magnitude of COVID-19 death risks is difficult to grasp, especially when population-level risks are small. Therefore, we converted the absolute risks of COVID-19 death into equivalents of death risk by a well-known, almost ubiquitous activity,^4^ driving/travelling by motor vehicle. We used estimates from the International Transport Forum Road Safety Annual Report 2018 for the number of road deaths per billion vehicle miles driven for each European country.^5^ For Spain, Italy, and Portugal there were only data available for number of road fatalities per 100,000 inhabitants. Since these tend to correlate reasonably well with road deaths per billion vehicle miles in Europe, we used for Italy and Portugal the same road deaths per billion miles as for Belgium, since Italy and Portugal have the same road deaths per 100,000 inhabitants as Belgium. Similarly, we used for Spain the same road deaths per billion miles as Germany. For USA locations, we used the state-specific data provided for 2018 by the Insurance Institute for Highway Safety.^6^ In other words, for each location we identified the distance that one has to travel by motor vehicle to expose oneself to the same hazard as the absolute COVID-19 death risk observed until May 1, 2020. We then divided the estimated miles travelled by the number of days that have passed since the first COVID-19 death was recorded in each location and until May 1, 2020. The result transforms the average risk of COVID-19 death per day during the period where COVID-19 deaths occur into an equivalent of miles travelled by car per day. The longer the distance, the higher the risk. Of note, for a typical death curve, e.g. as documented in Wuhan,^7^ the miles travelled per day estimate may be overestimated, if the covered fatality period extends beyond the peak of the curve, since the remaining days with COVID-19 deaths have fewer deaths per day than the already captured days that include the peak.

Finally, we sought information from the situational reports and from personal communications with the respective health authorities on how many COVID-19 deaths had been documented in people <65 years old who had no underlying predisposing conditions. Predisposing conditions for worse outcome in COVID-19 may include^8–10^ cardiovascular disease, hypertension, diabetes, chronic obstructive pulmonary disease and severe asthma, kidney failure, severe liver disease, immunodeficiency, and malignancy. We followed the data collection principles of each national and state organization on how underlying conditions were defined. Data were readily available in published reports or press releases for France,^11^ Georgia^12^ and New York City.^13^ We contacted all other national agencies and state departments of health when we could find contact information and thus we obtained additional such data according to the presence or not of underlying conditions from the Italian COVID-19 team (personal communication, Luigi Palmieri), from the Dutch COVID-19 team (personal communication, Susan van den Hof), and from the Swedish National Board of Health and Welfare (personal communication, Erik Wahlström).^14^ We encourage other organizations to send us similar data, as they become available, so that they can be incorporated in further updates. We avoided performing a formal meta-analysis, since these data are using different definitions of eligible comorbidities and data collection methods.

## RESULTS

### Eligible data

Eighteen countries (Belgium, Brazil, Canada, China, France, Germany, Iran, India, Ireland, Italy, Mexico, Netherlands, Portugal, Spain, Sweden, Switzerland, Turkey, United Kingdom) and 13 US states (California, Connecticut, Florida, Georgia, Illinois, Indiana, Louisiana, Maryland, Massachusetts, Michigan, New Jersey, New York, Pennsylvania) fulfilled the first eligibility criterion and of those, 12 countries (Belgium, Canada, France, Germany, Ireland, Italy, Netherlands, Portugal, Spain, Sweden, Switzerland, United Kingdom)^11,15–25^ as well as 12 states (California, Connecticut, Florida, Georgia, Illinois, Indiana, Louisiana, Maryland, Massachusetts, Michigan, New Jersey, New York)^12,26–36^ had some available data on required age categories.

### Death with age stratification

As shown in Table 1, individuals with age <65 accounted for only 4.8-9.3% of all deaths in 10 of the 11 European countries and in Canada, while they accounted for 13% of all deaths in the United Kingdom. Among the 12 US locations, the proportion of deaths contributed by individuals <65 ranged from 7.8-23.9% of all deaths. A lion’s share of all deaths are accounted by individuals 80 years or older in Europe (range, 54-69%) and Canada (67%), and the same appears to be true in the US, but with wider variability across states (range, 36-63%). Among all patients who died, men represented 49.7-63.3% of all deaths in different locations and there is a more prominent preponderance of men among patients who died younger than 65 (range, 59.1-77.6% of deaths <65) in locations where this information is available.

**Table 1.** Proportion of COVID-19 deaths contributed by specific age groups and proportion of men among these deaths.

| Location (date report) | Total deaths<sup>^</sup> | % of deaths with age <65 among total deaths | % of deaths with age ≥80 among total deaths | % male deaths (among all deaths) | % male deaths among deaths <65) |
| --- | --- | --- | --- | --- | --- |
| *Countries* |  |  |  |  |  |
| Belgium (Apr 23) | 6679 | 4.8 | 57.8** | 51.7 | 67.7 |
| Canada (Apr 23) | 863 | 5.3 | 66.5 | 51.1 | ND |
| France (Apr 21) | 7541 | 9.3 | 75.5** | 55.2 | ND |
| Germany (Apr 23) | 5321 | 4.3 [7.3]* | 63.1 | 57.0 | 74.7 [74.9]* |
| Ireland (Apr 21) | 789 | 8.4 | ND | ND | ND |
| Italy (Apr 23) | 23188 | 4.8 [8.5]* | 54.4 | 63.3 | 77.7 [77.6]* |
| Netherlands (Apr 24) | 4289 | 5.6 | 59.3 | 57.9 | 70.8 |
| Portugal (Apr 23) | 854 | 3.6 [6.6]* | 67.2 | 49.9 | 67.7 [67.9]* |
| Spain (Apr 23) | 13105 | 4.7 [7.7]* | 60.7 | 59.0 | 66.6 [68.1]* |
| Sweden (Apr 24) | 2152 | 5.1 [7.8]* | 63.4 | 56.4 | 76.3 [73.6]* |
| Switzerland (Apr 24) | 1309 | 2.5 [4.9]* | 69.2 | 58.7 | 69.7 [72.2]* |
| UK (Apr 10) | 11287 | 13.0 | 33.5*** | 59.7 | 63.1 |
| *US locations* |  |  |  |  |  |
| California (Apr 23) | 1562 | 23.0 | ND | 60.0 | ND |
| Connecticut (Apr 24) | 1764 | 7.3 [11.9]* | 56.2 | ND | ND |
| Florida (Apr 24) | 1046 | 17.4 | 59.3 | ND | ND |
| Georgia (Apr 24) | 899 | 23.9 | 35.7 | 54.4 | 59.1 |
| Illinois (Apr 24) | 1795 | 11.3 [17.1]* | 41.3 | 57.9 | ND |
| Indiana (Apr 23) | 741 | 9.5 [16.2]* | 43.2 | 56.0 | ND |
| Louisiana (Apr 24) | 1601 | 15.9 [22.2]* | ND | ND | ND |
| Maryland (Apr 24) | 622 | 12.7 [18.2]* | 42.9 | 51.6 | ND |
| Massachusetts (Apr 24) | 2556 | 4.6 [7.8]* | 63.4 | 49.7 | ND |
| Michigan (Apr 24) | 3085 | 14.0 [20.3]* | 39.0 | 55.0 | ND |
| New Jersey (Apr 24) | 5062 | 21.6 | 45.5 | ND | ND |
| New York (Apr 24) | 16162 | 15.4 [22.1]* | 37.6 | 59.6 | ND |

ND: no data; ^Using deaths' data with available information on age; \* data available only for the group with age <60 (the number shown in brackets is the approximated estimate for age <65 assuming that a third of the deaths in the 60-69 bracket are in <65 years old people, as suggested by other countries where data are available on 5-year age intervals); \*\* data available only on age $\geq 75$ \*\*\* data available only on age $\geq 85$

### Relative risk of dying with COVID-19 for individuals <65 years old versus older individuals at the population level

As shown in Table 2, the percentage of the population <65 years old varied from 75.18% in Italy to 86.31% in Illinois. People <65 years old had overall a 14- to 84-fold lower risk of COVID-19 death than those ≥65 years old; for continental European countries and Canada, individuals <65 years old had 36-to 84-fold lower risk of COVID-19 death than older individuals; while for the UK and US locations, the relative risk was somewhat smaller, with those <65 years old having 14- to 56-fold lower risk of dying than older individuals.

**Table 2.** Age distribution in the general population and relative risk of dying from COVID-19 for age ≥65 versus <65

| Location | Percentage of population <65 years in the general population (%) | Relative risk of COVID-19 death for those $\geq 65$ years versus those <65 years |
| --- | --- | --- |
| *Countries* |  |  |
| Belgium | 80.99% | 84 |
| Canada | 82.35% | 83 |
| France | 79.61% | 38 |
| Germany | 78.44% | 46 |
| Ireland | 85.78% | 66 |
| Italy | 76.99% | 36 |
| Netherlands | 80.39% | 69 |
| Portugal | 77.64% | 49 |
| Spain | 80.35% | 49 |
| Sweden | 79.80% | 47 |
| Switzerland | 81.16% | 84 |
| UK | 81.49% | 29 |
| *US locations* |  |  |
| California | 85.38% | 20 |
| Connecticut | 84.02% | 39 |
| Florida | 75.18% | 14 |
| Georgia | 83.32% | 16 |
| Illinois | 86.31% | 31 |
| Indiana | 83.63% | 26 |
| Louisiana | 85.82% | 21 |
| Maryland | 84.30% | 24 |
| Massachusetts | 82.57% | 56 |
| Michigan | 83.00% | 19 |
| New Jersey | 83.98% | 19 |
| New York | 85.03% | 20 |

As shown in Table 3, within the age group of <65, almost all deaths occurred in the range of 40-65 years. The group <40 corresponds to 52-63% of the age group <65, but only ≤1% of COVID-19 deaths occurred in people <40 years old in continental Europe and Canada. The proportion was a bit higher in most US locations and the United Kingdom.

**Table 3.** COVID-19 deaths in patients with age <40 years and proportion of people <40 years in the population

| Location | n (%) of COVID-19 | Percentage of | Percentage of population <40 |
| --- | --- | --- | --- |

|  | deaths with<br>age <40 | population<br><40 years in<br>the general<br>population<br>(%) | years among the<br>population <65<br>years (%) |
| --- | --- | --- | --- |
| *Countries* |  |  |  |
| Belgium | 20 (0.3%)* | 47.65% | 58.83% |
| Canada | 6 (0.7%) | 48.78% | 59.23% |
| France | 71 (0.9%)* | 47.42% | 59.57% |
| Germany | 8 (0.2%)** | 42.95% | 54.75% |
| Ireland | 13 (1.7%)* | 53.38% | 62.24% |
| Italy | 57 (0.2%) | 39.77% | 51.66% |
| Netherlands | 9 (0.2%) | 46.32% | 57.61% |
| Portugal | 0 (0.0%) | 41.23% | 53.10% |
| Spain | 80 (0.6%) | 42.44% | 52.81% |
| Sweden | 12 (0.6%) | 48.74% | 61.08% |
| Switzerland | 5 (0.4%) | 46.21% | 56.93% |
| UK | 133 (1.2%) | 49.56% | 60.82% |
| *US locations* |  |  |  |
| California | ND | 54.08% | 63.34% |
| Connecticut | 20 (1.1%) | 49.20% | 58.55% |
| Florida | 30 (2.9%)* | 44.67% | 59.42% |
| Georgia | 22 (2.4%) | 52.36% | 62.84% |
| Illinois | 38 (2.1%) | 53.40% | 61.87% |
| Indiana | 7 (1.0%) | 52.10% | 62.29% |
| Louisiana | 42 (2.6%) | 54.43% | 63.43% |
| Maryland | 16 (2.2%) | 51.12% | 60.64% |
| Massachusetts | 8 (0.3%) | 49.72% | 60.21% |
| Michigan | 31 (1.0%) | 49.95% | 60.18% |
| New Jersey | ND | 49.92% | 59.44% |
| New York | 301 (1.9%) | 52.08% | 61.25% |

ND: no data; \*Data shown for the group with age <45 years (not available for age <40 years) \*\*Data shown for the group with age <35 years (not available for age <40 years)

### Absolute risk of death with COVID-19 at the population level

Table 4 shows the estimates of the absolute risk of dying with COVID-19 at the population level for people <65 years old and for those ≥80 years old as of May 1. For these estimates we used the total number of deaths as of the close of day May 1, 2020, and not just those where age information was available (as in Table 1), assuming that the age stratification would be quite similar in all deaths as in the ones where age strata information has been released as of April 24. Also shown is the 7-day moving period that had the largest number of deaths. As shown, for most of the locations, the peak period was already many days, or even several weeks before May 1, suggesting that the epidemic fatality waves had already reached substantial maturity as of May 1. The waves of fatalities in Canada, Ireland, Maryland and possibly Illinois were the least complete. The absolute risk of death for people <65 years old ranged widely from 6 per million in Canada to 249 per million in New York. Connecticut, Louisiana, Michigan, New Jersey, and New York had an absolute risk of death exceeding 90 per million. Based on the stage of the epidemic waves in other locations, assuming a symmetric death curve, no other locations were likely to reach that level of absolute risk of death (≥90 per million) for people <65 years old, perhaps with the exception of the UK. The absolute risk of death for people ≥80 years old ranged from approximately 0.3 in a thousand in Florida to 10.6 in a thousand (~1 percent) in New York.

**Table 4.** COVID-19 deaths, peak 7-day period for deaths, population count and absolute risk of COVID-19 death for age groups <65 (per million) and ≥80 (per thousand)

| Location | Day of first documented COVID-19 death# | Peak 7-day COVID-19 death count as of May 1 | Total COVID-19 deaths as of May 1 (n)* | Total COVID-19 deaths in peak 7-day (n) | Population <65 years (n) | Population $\geq 80$ years (n) | Risk for people <65 (per million) | Risk for people $\geq 80$ (per thousand) |
| --- | --- | --- | --- | --- | --- | --- | --- | --- |
| *Countries* |  |  |  |  |  |  |  |  |
| Belgium | Mar 10 | Apr 7-13 | 7703 | 2132 | 9346151 | 658753 | 40 | ND |
| Canada | Mar 9 | Apr 25-May 1 | 3391 | 1089 | 30808712 | 1628779 | 6 | 1.4 |
| France | Feb 14 | Apr 2-8 | 24594 | 6837 | 51848623 | 4018291 | 44 | ND |
| Germany | Mar 9 | Apr 12-18 | 6481 | 1667 | 65508502 | 5737398 | 7 | 0.7 |
| Ireland | Mar 11 | Apr 19-25 | 995 | 492 | 4188062 | 151934 | 20 | ND |
| Italy | Feb 22 | Mar 27-Apr 2 | 26049 | 5700 | 46616108 | 4465708 | 47 | 3.2 |
| Netherlands | Mar 6 | Mar 30-Apr 5 | 4893 | 1120 | 13745168 | 819669 | 20 | 3.5 |
| Portugal | Mar 16 | Apr 7-13 | 1007 | 224 | 7939820 | 671048 | 8 | 1.0 |
| Spain | Feb 13 | Mar 28-Apr 3 | 24824 | 6060 | 37553712 | 2901252 | 51 | 5.2 |
| Sweden | Mar 10 | Apr 9-15 | 2653 | 674 | 8009176 | 522106 | 26 | 3.2 |
| Switzerland | Mar 5 | Mar 28-Apr 3 | 1435 | 352 | 6972924 | 448632 | 10 | 2.2 |
| UK | Feb 28 | Apr 7-13 | 27510 | 6602 | 55031155 | 3418559 | 65 | ND |
| *US locations* |  |  |  |  |  |  |  |  |
| California | Feb 6 | Apr 18-24 | 2073 | 579 | 34100000 | 1871089 | 14 | ND |
| Connecticut | Mar 17 | Apr 13-19 | 2339 | 492 | 2993800 | 143077 | 93 | 9.2 |
| Florida | Mar 6 | Apr 13-19 | 1314 | 316 | 16533700 | 2454485 | 14 | 0.3 |
| Georgia | Mar 12 | Apr 14-20 | 1167 | 254 | 8945200 | 721159 | 31 | 0.6 |
| Illinois | Mar 17 | Apr 24-30 | 2457 | 667 | 10926400 | 327782 | 38 | 3.1 |
| Indiana | Mar 15 | Apr 21-27 | 1062 | 260 | 5641400 | 353454 | 30 | 1.3 |
| Louisiana | Mar 14 | Apr 5-11 | 1927 | 438 | 3986700 | 137384 | 107 | ND |
| Maryland | Mar 17 | Apr 19-25 | 1098 | 344 | 5128000 | 290416 | 39 | 1.6 |
| Massachusetts | Mar 18 | Apr 18-24 | 3716 | 1178 | 5760800 | 426497 | 50 | 5.5 |
| Michigan | Mar 18 | Apr 10-16 | 3866 | 1017 | 8337500 | 486629 | 94 | 3.1 |
| New Jersey | Mar 11 | Apr 15-21 | 7538 | 1948 | 7505000 | 423174 | 217 | 8.1 |
| New York | Mar 14 | Apr 8-14 | 18610 | 5345 | 16529700 | 662769 | 249 | 10.6 |

Peak 7-day period of deaths is based on data as of May 1 and thus higher peaks and/or a second wave cannot be fully excluded. For France, Germany, Ireland, Italy, Portugal, Spain, Michigan, New Jersey, and New York only date of deaths reported were available, not actual date of death. ND: no data to allow calculation. # Based on official reports or news articles, but cannot exclude earlier COVID-19 deaths that went undetected \* updates are as of April 28 for Ireland, and April 30 for California and Indiana

Table 5 shows the conversion of the absolute risk of COVID-19 death as of May 1 into the equivalent death risk from motor vehicle travelled miles. The distances (corresponding to equivalent death risks) ranged from driving a total of 711 miles to 32739 miles. Dividing by the number of days since the first documented COVID-19 death, the average daily risk of COVID-19 death for an individual <65 years old in 11 of the 12 European countries or Canada is equivalent to driving between 13 and 101 miles per day during this period (47–79 days). California, Florida, Georgia, Illinois, Indiana, and Maryland are also in the same range of daily risk over 46–85 days. Conversely, the risk is higher in the UK and in the other 6 states in the USA (driving 143–668 miles per day) for the 45–64 days during which they have witnessed COVID-19 deaths.

**Table 5.** Absolute risk of COVID-19 death expressed as equivalent of death risk from associated with motor vehicle driving over given distances.

| Location | Road deaths per billion miles travelled | Risk of COVID-19 death for <65 year old people as total miles travelled equivalent (until May 1) | Days with COVID-19 deaths (as of May 1) | Risk of COVID-19 death for <65 year old people as miles travelled per day equivalent |
| --- | --- | --- | --- | --- |
| *Countries* |  |  |  |  |
| Belgium | 11.7 | 3381 | 53 | 64 |
| Canada | 8.2 | 711 | 54 | 13 |
| France | 9.3 | 4743 | 78 | 61 |
| Germany | 6.8 | 1062 | 54 | 20 |
| Ireland | 6.1 | 3272 | 49 | 67 |
| Italy | 11.7* | 4060 | 70 | 58 |
| Netherlands | 7.6 | 2623 | 57 | 46 |
| Portugal | 11.7* | 715 | 47 | 15 |
| Spain | 6.8* | 7485 | 79 | 95 |
| Sweden | 5.3 | 4875 | 53 | 92 |
| Switzerland | 5.1 | 1977 | 58 | 34 |
| UK | 5.5 | 11816 | 64 | 185 |
| *US locations* |  |  |  |  |
| California | 10.2 | 1371 | 85 | 16 |
| Connecticut | 9.3 | 9997 | 46 | 217 |
| Florida | 14.1 | 981 | 57 | 17 |
| Georgia | 11.4 | 2735 | 51 | 54 |
| Illinois | 9.6 | 4005 | 46 | 87 |
| Indiana | 10.5 | 2904 | 47 | 62 |
| Louisiana | 15.3 | 7013 | 49 | 143 |
| Maryland | 8.4 | 4639 | 46 | 101 |
| Massachusetts | 5.4 | 9317 | 45 | 207 |
| Michigan | 9.5 | 9908 | 45 | 220 |
| New Jersey | 7.3 | 29719 | 52 | 572 |
| New York | 7.6 | 32739 | 49 | 668 |

\*Approximation (see Methods, we welcome provision of any more precise estimates)

### COVID-19 deaths in individuals <65 years old without underlying conditions

Data on deaths in patients <65 years old without any underlying conditions (comorbidities) were available for France, Italy, Netherlands, Sweden, Georgia, and New York City. As shown in Table 6, the proportion of these deaths ranged from 0.6-2.6% of all COVID-19 deaths. The highest percentage was seen in France, based on deaths for which electronic death certificates were available as of April 21, and completeness of death certificate information is unknown.^11^ The second largest percentage was seen in Sweden, based on death counts that included also probable deaths without laboratory confirmation; moreover, only cardiovascular disease, hypertension, diabetes, and pulmonary disease were counted as comorbidities in this assessment, so it is possible that additional patients may have had other comorbidities.^14^

**Table 6.**
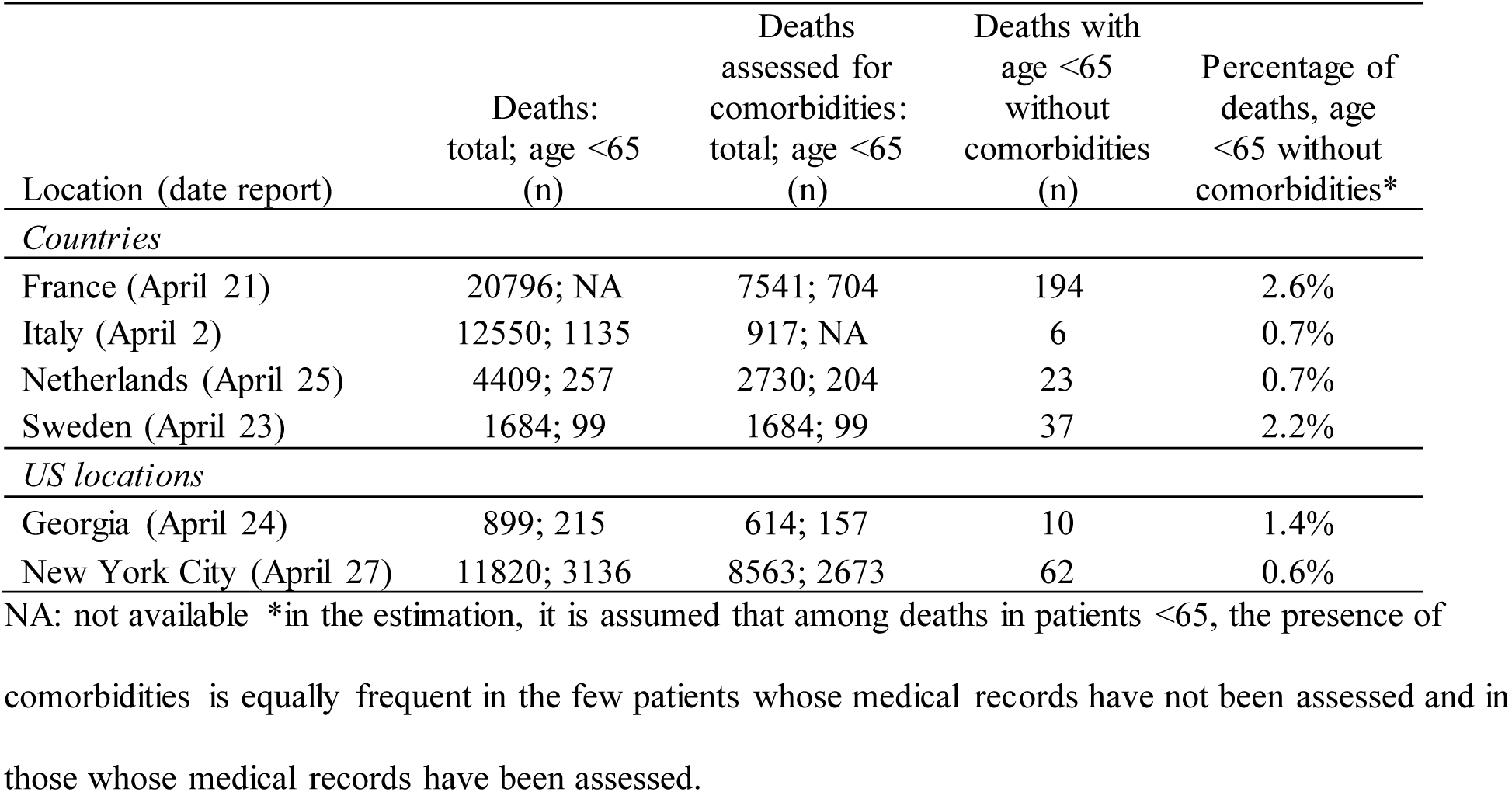
COVID-19 deaths in individuals <65 years old without underlying conditions

| Location (date report) | Deaths:<br>total; age <65<br>(n) | Deaths<br>assessed for<br>comorbidities:<br>total; age <65<br>(n) | Deaths with<br>age <65<br>without<br>comorbidities<br>(n) | Percentage of<br>deaths, age<br><65 without<br>comorbidities* |
| --- | --- | --- | --- | --- |
| *Countries* |  |  |  |  |
| France (April 21) | 20796; NA | 7541; 704 | 194 | 2.6% |
| Italy (April 2) | 12550; 1135 | 917; NA | 6 | 0.7% |
| Netherlands (April 25) | 4409; 257 | 2730; 204 | 23 | 0.7% |
| Sweden (April 23) | 1684; 99 | 1684; 99 | 37 | 2.2% |
| *US locations* |  |  |  |  |
| Georgia (April 24) | 899; 215 | 614; 157 | 10 | 1.4% |
| New York City (April 27) | 11820; 3136 | 8563; 2673 | 62 | 0.6% |

NA: not available \*in the estimation, it is assumed that among deaths in patients <65, the presence of
comorbidities is equally frequent in the few patients whose medical records have not been assessed and in those whose medical records have been assessed.

The lowest percentage (0.6% of all COVID-19 deaths) was seen in New York City, and it was based on assessment of comorbidities on approximately three quarters of the reported deaths. Of 3136 deaths at age <65 years, 2673 had been assessed for the presence of underlying diseases.

Similarly low percentages (0.7% of all COVID-19 deaths) were seen in Italy and Netherlands based on more detailed medical chart review. In Italy, this review is done on a subset of the deaths (n=917 as of April 2). In the Netherlands, information on underlying diseases is sought since April 10th only for those who died <70 years of age. Of 257 deaths at age <65 years, information on underlying diseases was available for 204. Finally, Georgia had an intermediate percentage (1.4% of all COVID-19 deaths), but relatively limited data.

## DISCUSSION

The evaluation of data from 12 countries and 12 US locations that are epicenters of the COVID-19 pandemic shows that non-elderly people <65 years old represent a very small fraction (4.8-9.3%) of all COVID-19 deaths in 11 European countries and Canada and between 7.8% and 23.9% of all COVID-19 deaths in 12 US locations, even though this age group represents the vast majority of the general population. Overall, the risk of death is 14-84-fold lower in non-elderly people <65 years old than in older individuals. The age-dependent risk gradient is modestly sharper in European countries and Canada versus most of the US locations. Regardless, the absolute risk of death in the non-elderly population is consistently very low even in these pandemic hotbeds. As of May 1, 2020, only 6 to 249 per million people in this age group have died with a COVID-19 diagnosis. Moreover, the vast majority of deaths in this age group occur in the age group 40-65 that comprises 37-48% of the population in the 0-65 years old bracket.

Of course, additional deaths may be recorded, as the epidemic wave progresses. However, it is likely that in all the locations that we examined, the peak daily deaths have already been reached (perhaps with the exception of Canada) and several countries have almost complete epidemic waves by May 1. Unless there is a further peak of deaths downstream, the total risk of death for the entire epidemic wave in these locations may not be much larger than what has been documented as of May 1, assuming a fairly symmetric epidemic wave, as in the case of Wuhan.^10^ In the absence of late surprises, it is very likely that when the epidemic wave completes its course, the risk of death in individuals <65 years old will remain very small.

For the whole COVID-19 fatality season to-date (starting with the date the first death was documented in each location), the average daily risk of dying from coronavirus for a person <65 years old is equivalent to the risk of dying driving a distance of 13 to 101 miles by car per day during that COVID-19 fatality season in 17 of the 24 hotbeds and 143-668 miles per day in the other 7 hotbeds (UK and 6 USA locations). For many hotbeds, the risk of death is in the same level roughly as dying from a car accident during daily commute. For example, the average commute is 31.5 miles per day for Americans according to the American Driving Survey^37^ and 44.2 miles per day round trip in Sweden.^38^ The highest daily risk of COVID-19 death (in New York) corresponds to a bit less than the risk of dying in a traffic accident while travelling daily from Manhattan to Rochester, NY round trip for these 49 days of COVID-19 fatalities-period. These per day risk estimates as of May 1 are a bit higher than those that we had estimated in the previous version of this work based on data as of April 4. However, given that the deaths have already peaked in most, if not all, examined locations, it is likely that the final per day estimated death risks when the waves mature will become again lower. People who are 40-65 years old may have about double that risk, while those 40 years old or younger have almost no risk at all of dying. Moreover, females may have 2-3 lower risk than males.

These risk estimates correspond to the main epicenters of the pandemic, since our eligibility criteria were set explicitly to include the locations with the highest numbers of deaths. Therefore, for the majority of countries around the world and for the majority of states and cities in the USA, the risk of death from COVID-19 this season for people <65 years old may have been even smaller than the risk of dying from a car accident during daily commute. We acknowledge that we cannot make any statement about the occurrence and magnitude of any second wave (e.g. in the fall/winter or next spring), but even for influenza the magnitude of the 2020-2021 wave is largely unpredictable.

We should also acknowledge that all the epicenters considered in this analysis are high-income countries with generally high life expectancy. For lower income countries with lower life expectancy, the proportion of deaths among younger age strata may be larger. For example, in India, life expectancy is almost a decade less than in the USA and almost 15 years less than in Switzerland, making octogenarians and nonagenarians few in relative terms. Not surprisingly, preliminary data suggest that 14% of COVID-19 deaths in India are in people below 45 years and another 34% in people 45-60.^39^ However, the overall population level death risk across all age groups (0.9 per million as of May 1, 2020) is much lower compared with the epicenters we analyzed; thus the absolute population risk of death for non-elderly individuals in India would be extremely low.

Some caveats about the data need to be discussed. Even though mortality is an unambiguous endpoint, attribution of death to a specific cause is often challenging and definitions of “COVID-19 death” vary across countries and sometimes even change within countries over time. For example, the presented age-stratified data on Canada and UK do not seem to include deaths that happened outside the hospitals. Such deaths were added to the death counts in the UK on April 28, 2020, but no available data on age-strata were available as of finalizing this paper on May 1, 2020. Different countries and US locations differ on the threshold of including deaths at care homes. For example, in Belgium, 53% of deaths come from care homes, but 94% of them have not had laboratory confirmation.^40^ New York City and some other US locations have also started counting in more recent counts also “probable” deaths without any COVID-19 laboratory confirmation, a debatable practice at best. Overall, some COVID-19 deaths may be missed, and others may be overcounted. Different death tallies are derived by different sites. For example, compilation of death certificate data from CDC had 37,308 confirmed or presumed deaths in the USA until the week ending April 25 (https://www.cdc.gov/nchs/nvss/vsrr/covid19/index.htm), while the popular Worldometer site had counted 54,256 confirmed or probable deaths as of the same date. Arbitration and proper calibration of death counts may need to await careful, in depth medical chart review and autopsy efforts. Under- and over-counting may be responsible for some of the heterogeneity that we observe in both relative and absolute risk estimates.

We should also acknowledge that we focused on mortality risk and not on hospitalizations. Empirical experience shows that COVID-19 has the potential to overwhelm specific hospitals, especially in settings where hospitals run close to maximum capacity even under regular circumstances, and when they serve high risk populations in cities with high population density and major congregations in mass events. Therefore, hospital preparedness is totally essential, regardless of whether the risk of death is very low in the general population. Similarly, work modeling hospital bed needs is useful. However, for understanding the risk of individuals from the general population, the analogy against deaths by motor vehicle accidents is still relevant, since motor vehicle accidents also result in many more people who require hospitalizations and who suffer major injuries beyond the numbers of those who die.

The death risk estimates that we calculated may also be corroborated by the perusal of patterns of excess mortality in the general population during the period of COVID-19 fatalities. For example, data from 24 European countries^41^ show that in the 6 weeks between week 10 and week 16 in 2020, the excess of deaths increased by 8146 deaths in the <65 years age category and by 107,343 deaths in the older age category. Therefore, the excess deaths in the <65 years category accounted for 7.1% of the total excess, a number very compatible with the 4.8-9.3% figure observed in 10 of the 11 European countries that we analyzed. However, one should be cautious in this comparison, because changes in other causes of deaths may also affect the magnitude of the overall excess during these 6 weeks. In fact, there is concern that COVID-19 measures may have taken a toll on other causes of death, e.g. people with heart attacks may fear for coming to hospitals for treatment.^42,43^

The large majority of the deaths in non-elderly individuals occur in patients who have underlying diseases. Based on existing data to-date,^8–10^ cardiovascular disease, hypertension, chronic obstructive pulmonary disease and severe asthma, diabetes, kidney failure, severe liver disease, immunodeficiency, and malignancy may confer an increased risk of adverse outcome. Individuals with these diseases should consider that their risk may be higher than average and rigorous prognostic models need to be developed to estimate with accuracy this increased risk. In non-elderly populations, the more prevalent of these conditions is cardiovascular disease and hypertension, with prevalence of approximately 10% in the 20-39 age group and 38% in the 40-59 age group in the USA^44^ and similarly high percentages in many other countries. We encourage public health authorities to start reporting systematically data separately on each of the major comorbidities according to age strata. Some data are available for the prevalence of these conditions across all age groups of COVID-19 deaths. For example, in the Netherlands, among COVID-19 deaths in people <70 years old, 39% had cardiovascular disease or hypertension, and 22% had chronic pulmonary disease. Comparing with the prevalence of these diseases in the general population,^45^ it is likely that ~2-fold increases in death risk may be reasonable to expect for people with these conditions in the general population. If so, the risk may remain very low, except in a minority of patients with the most severe manifestations of the underlying diseases.

We could retrieve data from 6 locations on the COVID-19 mortality of people who were both <65 years old and had no underlying diseases. Consistently, the data suggest that these deaths are remarkably uncommon, although their exact percentage contribution to all COVID-19 deaths varies modestly across locations (0.6-2.6%). It is likely that much of this variability may reflect differences on how underlying conditions are captured. We also caution that some people with no recorded comorbidities may have had some underlying diseases, but these where not reported in a crisis setting, or these conditions may have been undiagnosed. Overall, this further strengthens the notion that for healthy non-elderly people, the risk of dying from COVID-19 this season has been infinitesimally small. This is in stark contrast with many news stories that focus on the demise of young people and the panic and horror that these widely reverberated stories are causing. It is very important for authorities in all countries and US locations to report carefully curated data on comorbidities and related death rates.

Another interesting observation is the higher share of deaths in the <65 years old group in most of the US locations as opposed to the European countries. The difference based on the current data is less prominent than what had been observed in our original analysis which had employed data until April 4. It is unknown whether the difference may shrink further as the US epidemic curves mature further. This pattern requires further investigation, but it may reflect unfavorable socioeconomic circumstances for victims of COVID-19 in the USA. It is important to study in more detail the socioeconomic profile of the COVID-19 victims, but preliminary data show that deaths cluster in areas with high levels of poverty and underprivileged populations and ethnic/racial minorities are over-represented among the victims.^46^ Some mostly low-wage occupations, including essential jobs, also may be prone to more exposure risk than other jobs where working remotely is feasible. COVID-19 may thus be yet another disease with a profile dependent on inequalities and generating even more inequalities. Of interest, influenza deaths seem to have a similar difference in age distribution between the USA and European countries like Italy: a larger proportion of influenza deaths in the USA tend to be in the <65 age group,^47^ as compared with Italy.^48^ Of course, a major difference between influenza and COVID-19 is that the latter does not cause deaths in otherwise healthy children, in contrast to influenza.

The vast majority of victims from COVID-19 are elderly people and in all European countries analyzed as well as Canada and most US locations, more than half and up to three quarters are at least 80 years old. The median age of death for COVID-19 tends to be similar or slightly smaller than the life expectancy of the population in each respective location. In several locations, large clusters of deaths come from nursing home facilities. Data from European countries suggest that 42-57% of all deaths have happened in care homes^49^ and many deaths in the US have also occurred in nursing homes.^38^ Moreover, the differentiation between dying “with” SARS-CoV-2 versus dying “from” SARS-CoV-2 may be difficult to make, and the vast majority of patients with COVID-19 have comorbidities and these could also contribute to the fatal outcome or may be even more important than SARS-CoV-2 in causing the death.^50^ Nursing homes and hospitalized patients (nosocomial infection) appear to account for a lion’s share of COVID-19 mortality. Overall, the loss of quality-adjusted life-years from COVID-19 may be much smaller than a crude reading of the number of deaths might suggest, once these features are accounted for.

The data that we have compiled allow to estimate also absolute risks of death in the highest risk group, i.e. elderly individuals ≥80 years old in these hot epicenters of the pandemic. These are markedly higher than the risks of death in individuals <65 years old. However, the absolute risk of death even in this highest age category to-date barely reach ~1% in the most hit location and in several locations it is lower than 1 in a thousand. Nevertheless, these risks are clearly high enough to warrant high alert. They suggest that, no matter what strategy is selected for addressing COVID-19 in the current or future epidemic waves should include special emphasis in protecting very elderly individuals.

As the data from the first epidemic wave of COVID-19 mature, knowledge of relative and absolute risks for different population strata are instrumental for carefully choosing next steps. Lockdowns have been implemented in many countries and this was a fully justified initial “better safe than sorry” approach in the absence of good data. However, long-term lockdowns may have major adverse consequences for health (suicides, domestic violence, worsening mental health, cardiovascular disease, loss of health insurance from unemployment, and famine, to name a few) and society at large.^41,51–54^ Lockdowns originally aimed to save the health care system, but as patients avoid coming to the hospitals for common, treatable problems,^55^ deaths may rise from non-treatment; also hospitals face an emerging crisis also leading to their demise. It is argued that lockdowns may be even harmful as a response to COVID-19 itself, if they broaden rather than flatten the epidemic curve.^56^ Information from seroprevalence and universal screening studies suggest that the frequency of infections is much larger than the documented cases and thus the overall infection fatality rate is much lower than previously thought.^57–59^ It seems that the vast majority of infections are either asymptomatic or mildly symptomatic and thus do not come to medical attention.^60^ These data also suggest that the infection fatality rate may be close to that of a severe influenza season (<0.2%) when the health system does not collapse and when massive nosocomial infections and nursing home spread are averted. Conversely, high infection fatality rates are seen when hospitals are overrun, and there are massive death loads from nosocomial-infected hospitalized patients and nursing home residents (e.g. in New York). Therefore, the finding of very low risk in the vast majority of the general population has major implications for strategic next steps in managing the COVID-19 pandemic. Tailored measures that maintain social life and the economy functional to avoid potentially even deaths from socioeconomic disruption, plus effective protection of select high-risk individuals (in particular in hospitals and nursing homes) may be a sensible option. Draconian measures of hygiene and infection control and universal, repeated testing of personnel in hospital and care facilities may achieve drastic reductions in deaths. Concurrently, the vast majority of the population may be reassured that their risks are very low.

## Data Availability

All data used for this manuscript will be shared upon request.

## Acknowledgments

We are grateful to Luigi Palmieri, Susan van der Hof and Erik Wahlström for providing additional data clarifications. We welcome provision of additional data and detailed information and any relevant data clarifications from all countries and states. As further information accumulates, this will allow updating and improving the volume of data and the precision of the estimates in the present analysis.

## Data sharing

All data used in this manuscript will be shared upon request.

